# What Does ’Polysubstance’ Really Mean? Comparing Drug-Involved Deaths in Los Angeles in CDC Records vs. Detailed Medical Examiner Data

**DOI:** 10.1101/2025.06.27.25330444

**Authors:** Joseph Friedman, Ruby Romero, Arthur Funnell, David Goodman-Meza, Chelsea L. Shover

## Abstract

**Introduction:** The ongoing overdose crisis in the United States is increasingly characterized by polysubstance involvement, shifting beyond single-drug fatalities to complex combinations that present novel public health challenges. However, most epidemiological literature describing the US overdose crisis has still primarily relied on traditional data sources, especially the CDC WONDER system that is limited to assessing drugs with established ICD-10 codes using a maximum of n=2 ICD-10 codes. In this analysis we leverage complete medical examiner records Los Angeles County examine the limitations of CDC WONDER data for detecting polysubstance drug deaths.

**Methods:** Deaths counts describing drug-involved deaths occurring in Los Angeles County were described using the CDC WONDER system, and medical examiner data from Los Angeles County. The average number of drugs, and the proportion of polysubstance deaths (involving 2 or more) involving 3 or more substances was summarized by year. Head-to-head comparisons were conducted of single-drug involvement, and UpSet plot visualization was employed for complex set analysis.

**Results:** The average number of drugs present per drug-involved death increased from 1.72 in 2012 to a peak of 2.29 in 2023. The percentage of polysubstance deaths (2+ substances) that would not be fully characterizable using the CDC WONDER system given a 2 drug limit), increased from 51% in 2012 to a peak of 74% in 2023. The total number of unique polysubstance combinations with >2 substances increased from n=11 in 2012 to n=57 in 2023 and was n=39 in 2024. Overall concordance in single-substance death counts between CDC WONDER and medical examiner records was high among n=6 substances of interest. Across n=42 drug and year pairs observed between 2018 and 2024, the overall R^2^ according to a Pearson correlation analysis was 0.99.

**Discussion:** Although concordance is very high for trends describing single-substance drug deaths, CDC WONDER fails to capture the majority of polysubstance deaths adequately. We find that a very large fraction of polysubstance deaths (about three quarters) are incompletely described using the 2 drug limit employed by CDC WONDER. This limitation has increased over time, as the complexity of polysubstance overdose deaths has increased. We illustrate a huge variety of polysubstance deaths that can be seen in medical examiner deaths. As the overdose crisis grows increasingly polysubstance in nature, improving the epidemiological tracking of deaths involving multiple drugs, and drugs not captured by ICD-10 codes currently, is of paramount importance. Here we highlight the gaps in existing traditional epidemiological sources which may serve as a motivating example for future improvements.

## Introduction

The ongoing overdose crisis in the United States is increasingly characterized by polysubstance involvement, shifting beyond single-drug fatalities to complex combinations that present novel public health challenges[1–4]. For instance, the fraction of U.S. overdose deaths involving both fentanyl and stimulants surged from 0.6% in 2010 to 32.3% in 2021[2]. This general shift towards polysubstance deaths is referred to as the "Fourth Wave" of the US overdose crisis [1].

The Fourth Wave of the overdose crisis increasingly features complex polysubstance combinations of various drugs like fentanyl and methamphetamine with more novel substances such as xylazine, medetomidine, nitazenes, novel synthetic benzodiazepines, synthetic cannabinoids, and others[5–8]. These polysubstance formulations are increasingly described using community-based drug checking technologies leveraging mass spectrometry, Fourier-transform infrared spectrometry, and other techniques[9].

However, most epidemiological literature describing the US overdose crisis has still primarily relied on traditional data sources – especially the CDC’s Wide-ranging Online Data for Epidemiologic Research (WONDER) system[2,10]. The CDC WONDER system is broadly used by researchers and policymakers to generate summary statistics on-demand, and it has many advantages, including being publicly available and producing continuously updated overdose statistics on an approximately 7 month lag (due to inherent lags in toxicology testing[11]).

Disadvantages of CDC WONDER include that it can only pull drug-involved deaths using established ICD-10 codes (e.g. the code T40.5 which codes for “Cocaine”). Many common substances, including fentanyl and methamphetamine, have no bespoke code, but are coded for using umbrella codes that have come to represent them de-facto (T40.4 “Other synthetic narcotics” and T43.6 “Psychostimulants with abuse potential”, respectively, for these two substances). Emerging evidence suggests that the veterinary sedative xylazine may now be commonly represented by the combination of T42.7 (“Antiepileptic and sedative-hypnotic drugs, unspecified”) or T46.5 (“Other antihypertensive drugs, not elsewhere classified”)[12].

Given that CDC WONDER does not facilitate any access to the free text fields on death certificates, there is no ability to systematically leverage information about drug-involved deaths with substances implicated that do not have an established ICD-10 code.

Another critical limitation of using CDC WONDER for tracking polysubstance deaths is that the platform allows for assessing deaths using a maximum of n=2 ICD-10 codes. A researcher can therefore generate counts of deaths involving two specific substances (e.g. involving xylazine and fentanyl) but not specifically describe groups of deaths involving 3+ substances (e.g. xylazine, fentanyl, and cocaine), or deaths involving 2 substances but specifically excluding a third substance (e.g. xylazine and cocaine, but not fentanyl).

CDC WONDER, in effect, aggregates data from the over 2,000 medical examiner and coroner’s jurisdictions across the US. An alternative strategy to leveraging CDC WONDER to track overdose deaths, is directly obtaining data from these medical examiners and coroners, and aggregating them leveraging fully detailed text fields described all drugs discovered using toxicological analysis[4]. We have used this approach across several jurisdictions to provide more in-depth analyses[4,5]. However considerable uncertainty exists regarding the degree to which CDC WONDER and medical examiner data perfectly represent one another – given differences in underlying data definitions. Furthermore, a large, but generally unquantified, fraction of overdose deaths involves >2 substances. We hypothesize that the fraction of deaths involving greater than 2 substances, and therefore incompletely characterized using CDC WONDER, is likely increasing over time.

In this analysis we leverage complete medical examiner records from the medical examiner’s office of Los Angeles County – the nation’s largest county, and likely one of the largest medical examiner jurisdictions. We compare CDC WONDER data for Los Angeles County to this more direct data source, to see which fraction of deaths are incompletely described using traditional approaches.

## Methods

Deaths counts describing drug-involved deaths occurring in Los Angeles County were described using the CDC WONDER system[10]. To mimic the nature of the medical examiner data, underlying causes of death were not restricted. Drug-involved deaths were pulled using multiple cause of death codes for fentanyl (T40.4), methamphetamine (T43.6), cocaine (T40.5), heroin (T40.1), prescription opioids (T40.2 and T40.3), and benzodiazepines (T42.4). Deaths were pulled by year and single substance involvement.

Medical examiner data from Los Angeles County were requested and assembled using previously described processes[4]. Specific substance involvement was determined searching across the various cause of death and other free text fields available from each death certificate, using natural language processing based models to classify these texts into different overdose types (published separately [13] and see code in Github repository: https://github.com/ROSLA-UCLA/ROSLA-HEAL-Data). Deaths were categorized as involving fentanyl, methamphetamine, cocaine, heroin, prescription opioids, benzodiazepines, alcohol, xylazine, and ‘others’. The ‘others’ category captured a large number of other substances that appeared less commonly.

The average number of drugs, and the proportion of polysubstance deaths (involving 2 or more) involving 3 or more substances was summarized by year. Given the use of the ‘other’ category, these summary statistics may represent a slight underestimate, or a trimmed mean, but there were relatively few deaths involving the ‘others’ category and only one other drug, therefore this limitation was not substantial.

All polysubstance combinations in medical examiner data containing more than 2 substances were visualized using UpSet plot visualization[14]. Data were visualized using R version 4.4.1 and the ggplot2 package. Study protocols were approved by the UCLA Institutional Review Board (IRB-22-1273).

## Results

The distribution of number of substances involved in drug involved deaths was visualized by year (Figure 1, Part A). The average number of drugs present per drug-involved death, and the percent of polysubstance deaths (2+ substances) that would not be fully characterizable using the CDC WONDER system given a 2 drug limit), were summarized by year (Figure 1, Part B and C). Over time, the fraction of deaths involving only one substance shrank, while the fraction containing 3 or more deaths increased (Figure 1, Part A). The average number of drugs present per drug-involved death increased from 1.72 in 2012 to a peak of 2.29 in 2023 (Figure 1, Part B). The percentage of polysubstance deaths (2+ substances) that would not be fully characterizable using the CDC WONDER system given a 2 drug limit), increased from 51% in 2012 to a peak of 74% in 2023.

**Figure 1.**
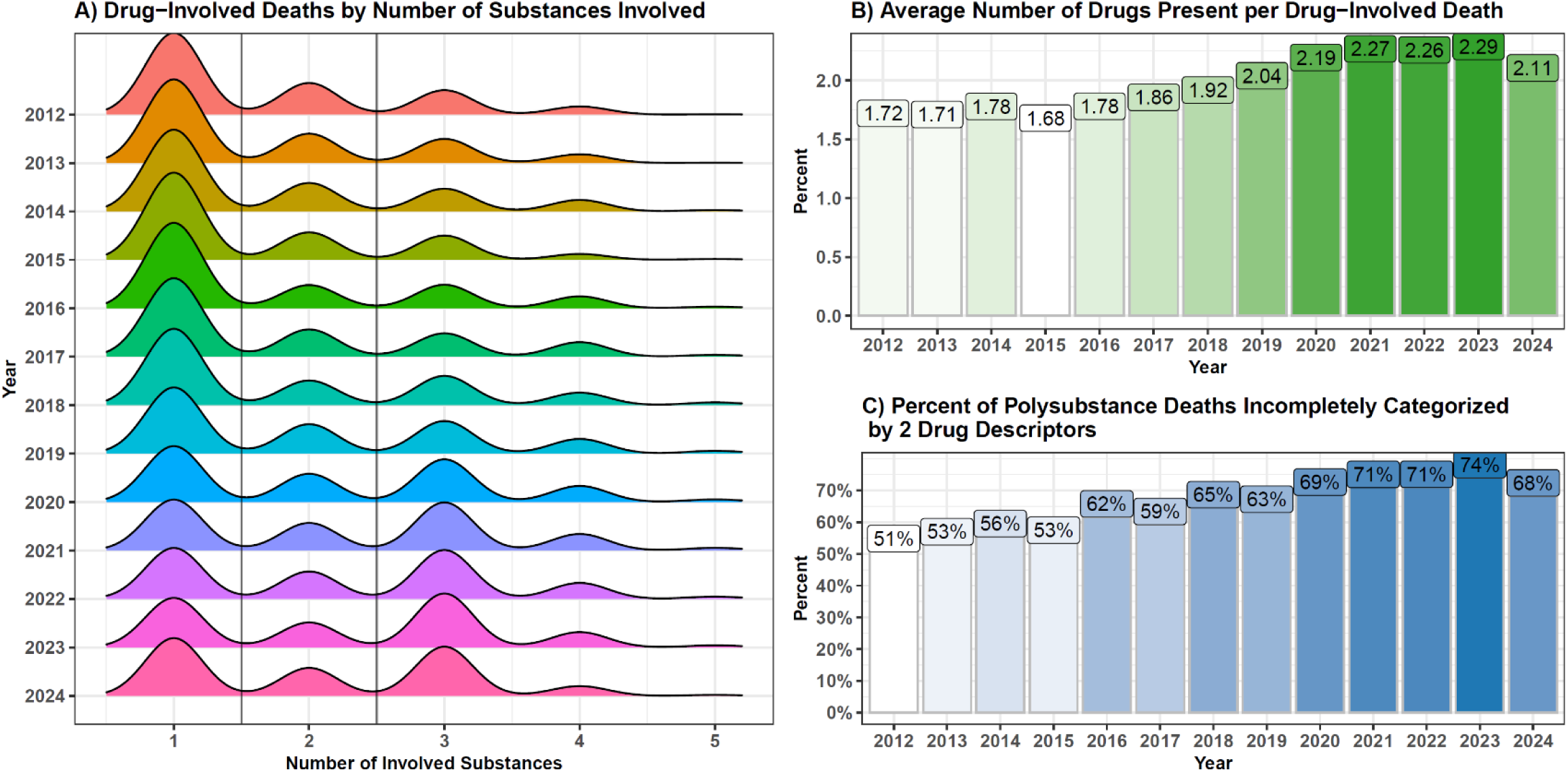
Description of the number of substances involved in drug-involved deaths in Los Angeles in Medical Examiner Data Part A) Shows the distribution of number of substances involved in drug involved deaths by year. Vertical lines separate 1 death and 2 deaths (the border of polysubstance vs non polysubstance) as well as 2 deaths and 3 deaths (the border between polysubstance deaths fully characterizable, or not fully characterizable, using the current CDC WONDER system). Part B) Average number of drugs present per drug-involved death by year. Intensity of color represents the magnitude of the number. Part C) Percent of polysubstance deaths (2+ substances) that would not be fully characterizable using the CDC WONDER system given a 2 drug limit) by year. Intensity of color represents the magnitude of the percent.

**Figure 2.**
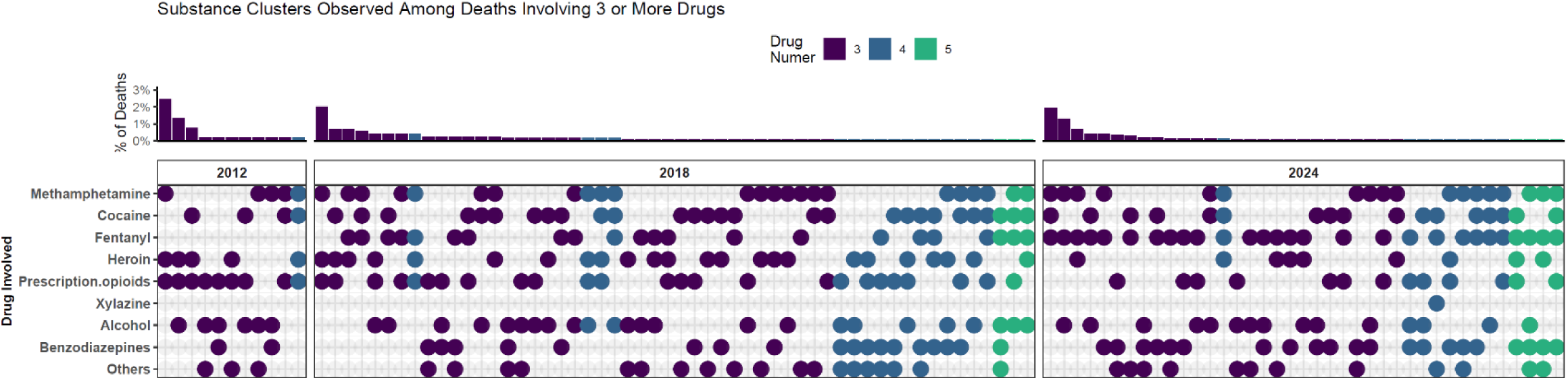
Substance Clusters Observed Among Deaths Involving 3+ Deaths Clusters of deaths involving 3 or more substances (which therefore could not be completely captured by CDC WONDER data) are shown for 2012, 2018, and 2024. One column represents a particular cluster of substances for a given year. On the bottom panel, one row represents the presence or absence (with a present or absent dot) of a given substance. Dot color corresponds to number of drugs involved. On the top panel, the proportion of all drug deaths in a given year represented by a given cluster is shown.

**Figure 3.**
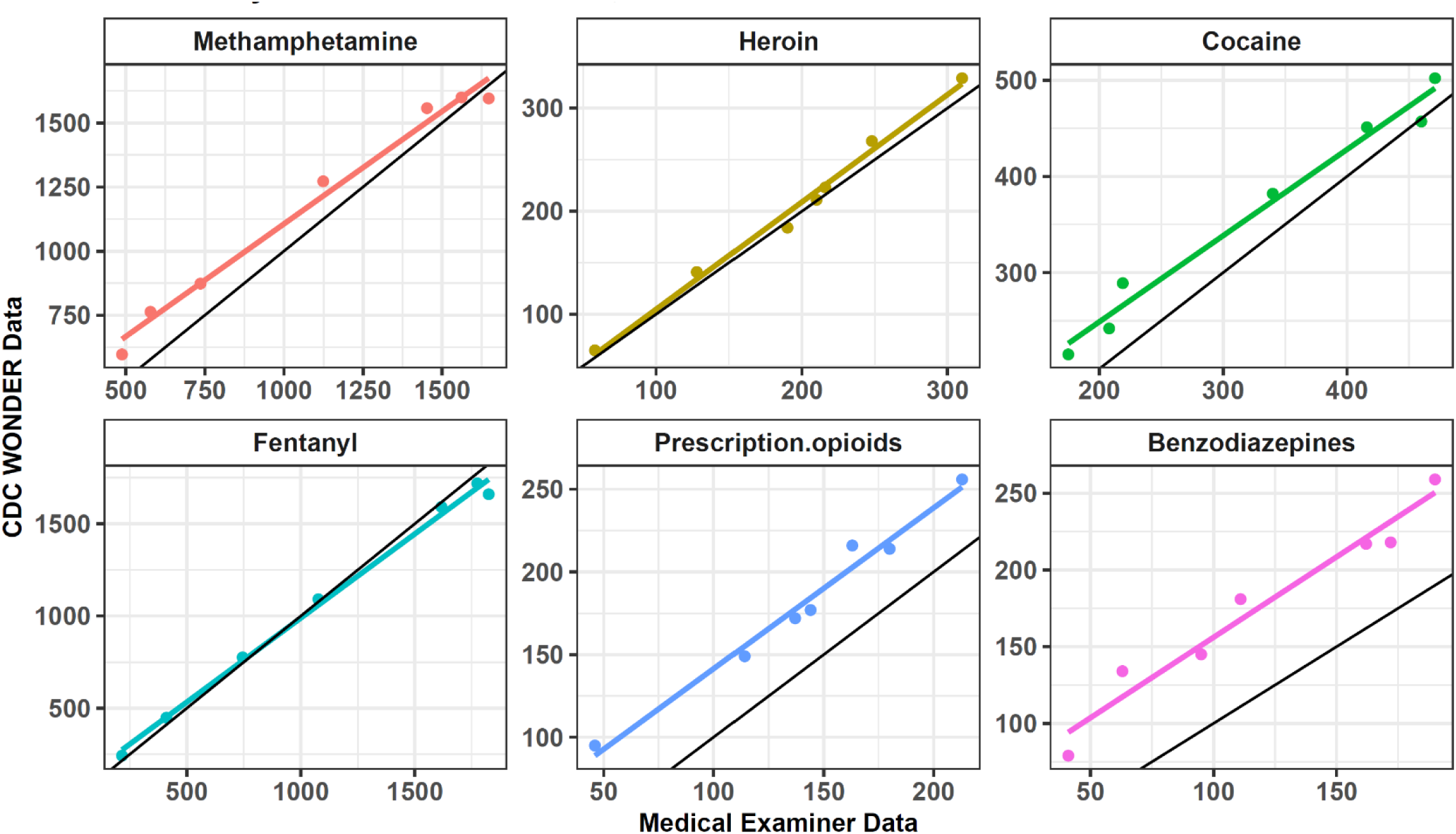
Comparing Counts of Drug-Involved Deaths in CDC Wonder vs. LA County Medical Examiner Data, 2018-2024 For 6 drugs of interest, counts of drug-involved deaths from CDC WONDER for LA County are compared on the y axis, to counts from medical examiner data for the same substance-involved deaths on the x-axis. A line of equality is shown for each substance in black. One point represents one year of data. 2024 includes on data through the end of August for both sources.

The full set of polysubstance deaths containing clusters of 3 or more substances (which therefore could not be completely captured by CDC WONDER data) were visualized for 2012, 2018, and 2024. The total number of unique polysubstance combinations with >2 substances increased from n=11 in 2012 to n=57 in 2023, and was n=39 in 2024 (of note this represents only 8 months of the year). The most prevalent cluster in 2012 was the combination of methamphetamine, heroin, and prescription opioids. The most commn cluster in 2024 was methamphetamine, cocaine, and fentanyl. In 2012 there were no combinations involving 5 drugs, and only n=1 combination involving 4 drugs. By 2024, n=9 involved 4 drugs, and n=4 involved 5 drugs.

Concordance in single-substance death counts were also assessed between CDC WONDER and medical examiner records, for 6 substances of interest. The overall concordance was high.

Across n=42 drug and year pairs observed between 2018 and 2024, the overall R2 according to a Pearson correlation analysis was 0.99. Fentanyl and heroin deaths had near perfect concordance. Methamphetamine death numbers were initially higher in CDC WONDER data in 2018 compared to medical examiner data for the same year, but concordance improved by 2023, with 1,599 and 1,561 deaths reported respectively. Deaths reported for prescription opioids, and benzodiazepines were higher among CDC WONDER compared to medical examiner data across all years examined.

## Discussion

This study leveraged complete medical examiner data from Los Angeles County (the nation’s most populous county) and compared it to CDC WONDER data to see which fraction of deaths are incompletely described using traditional approaches. Although concordance is very high for trends describing single-substance drug deaths, CDC WONDER fails to capture the majority of polysubstance deaths adequately. We find that a very large fraction of polysubstance deaths (about three quarters) were incompletely described using the 2 drug limit employed by CDC WONDER. This limitation increased over time, as the complexity of polysubstance overdose deaths increased. We illustrate a huge variety of polysubstance deaths that can be seen in medical examiner deaths. Much of this variation was lost in the national aggregation process that underpins CDC WONDER and other traditional epidemiological approaches.

Some solutions to the limitations of CDC WONDER have already been proposed. For instance, the State Unintentional Drug Overdose Reporting System (SUDORS) system aggregates more detailed death records from the majority of states[15], in a similar fashion to independent research groups that have aggregated medical examiner records[4]. Although SUDORS has a public data visualization providing some summary statistics, the underlying data are not available to researchers or the public, limiting the usefulness of the information. More detailed text information is also collected by the National Vistal Statistics System (which also feeds mortality data in CDC WONDER), which has been used for specific analyses conducted by government researchers[16]. Therefore, there is no clear logistical reason why CDC WONDER or another platform could not provide a) aggregate statistics using more than two multiple cause of death codes allowing for more complex polysubstance death assessments and b) access to information contained in free text fields allowing for the quantification of substances not captured in existing ICD-10 codes. Investing in these practices would likely greatly improve the productivity of research in this area. However, it would require government investment in transparency, and updates to existing systems.

As the overdose crisis grows increasingly polysubstance in nature, improving the epidemiological tracking of deaths involving multiple drugs, and drugs not captured by ICD-10 codes, is of paramount importance. Here we highlight the gaps in existing traditional epidemiological sources which may serve as a motivating example for future improvements.

## Data Availability

The CDC data used in this study are available at: https://wonder.cdc.gov/
The medical examiner data used in this study cannot be shared by the authors, but summary statistics may be shared by the authors upon reasonable request.

https://github.com/ROSLA-UCLA/ROSLA-HEAL-Data

